# Vivaray Hb pro noninvasive hemoglobin device: a prospective diagnostic accuracy study measuring agreement with a calibrated blood cell counter

**DOI:** 10.64898/2026.06.29.26356869

**Authors:** Devesh Kumar, Sunita Kapoor, Ambanna Gowda, Dipti Gupta, Harsh Mittal, Suneet Sood

**Affiliations:** IR Innovate Research Pvt. Ltd., Noida, Uttar Pradesh, India; City X-Ray and Scan Clinic Pvt. Ltd., 5A/34, Tilak Nagar, New Delhi, India; Citizen Hospital, Dispensary Road, Kalasipalya, Benagluru, India; Dr. Dipti’s Multispeciality Clinics, Rajendra Nagar, Sahibabad, UP, India; Panchsheel Hospital, New Delhi, India; Department of Surgery, IMU University, Malaysia

**Keywords:** Anemia, screening, oximetry, transcutaneous, point of care testing, photoplethysmography

## Abstract

**Background:** Non-invasive hemoglobin measurement offers a painless and rapid alternative to conventional blood-based testing. The Vivaray Hb pro is a handheld photoplethysmography-based device designed for point-of-care hemoglobin assessment without blood sampling. We evaluated the clinical performance of the Vivaray Hb pro by comparing device-generated hemoglobin values with those obtained from a calibrated laboratory blood cell counter.

**Methods:** In this cross-sectional, non-randomized clinical performance study, participants aged ≥8 years were prospectively recruited. Hemoglobin was measured non-invasively using the Vivaray Hb pro and compared with venous blood samples analyzed on a calibrated Coulter counter. Agreement between methods was assessed using Bland–Altman analysis, including regression-based evaluation for proportional bias. Mean absolute error (MAE) and proportions of measurements within tolerance limits were also calculated. Complete paired measurements were available for 763 individuals.

**Results:** Bland–Altman analysis demonstrated hemoglobin-dependent bias, with overestimation at lower hemoglobin levels and underestimation at higher levels. Regression-based analysis showed proportional bias (β_1_ = −0.178), indicating decreasing difference with increasing hemoglobin concentration. The MAE was 1.5 g/dL but was lower (1.2) in the clinically predominant ranges of 8.1-13 g/dl.

**Comment:** The results support the use of the Vivaray Hb pro as a noninvasive hemoglobin screening and point-of-care assessment tool, particularly in settings where rapid, painless, and repeatable measurements are desirable.

## Introduction

Anemia remains a major health problem, affecting about 30% of women of reproductive age, 35% of pregnant women, and 40% of children aged 6–59 months worldwide.(1) Prevalence is higher in low- and middle-income countries, where nutritional deficiencies and infections are more common.

Screening is recommended for populations at higher risk of anemia, including pregnant women, young children, women of reproductive age, and individuals with chronic illnesses or nutritional deficiencies.(2) Early detection in pregnancy helps prevent adverse maternal and child outcomes and supports timely intervention. Global health agencies emphasize routine screening in regions with high anemia prevalence, particularly in resource-limited settings.

A non-invasive, point-of-care hemoglobin device could greatly simplify anemia screening by eliminating the need for finger-prick or venous blood sampling. Invasive methods cause discomfort and have small but measurable risks, (3) and non-invasive methods are likely to be more acceptable among children and pregnant women. They also facilitate rapid, on-site testing in community clinics, and enable continuous monitoring where needed.(4)

### Non-invasive hemoglobin devices

Non-invasive hemoglobin-measuring devices have evolved substantially over the past two decades, driven by the need for rapid, painless, and point-of-care assessment of anemia in both clinical and community settings.

While accuracy varies across devices and populations, the overall trend shows steady improvement, supporting the role of non-invasive technologies in anemia screening especially I children, chronic disease management, trauma care and perioperative monitoring.(5,6)

The principles of non-invasive hemoglobin estimation are an extension of those of pulse oximetry. Light passed through the finger is absorbed depending on the constituents of blood, especially hemoglobin. During systole, the vessels in the finger fill up, there is more hemoglobin to absorb the light. This situation is reversed during diastole. In this way the pulse oximeter is able to detect the heart rate. Per the Beer-Lambert Law, the concentration of a given solute in a solvent can be determined by the amount of light that is absorbed by the solute at a specific wavelength.(7)

Oxyhemoglobin absorbs near-infrared light at 940 nm and at 577 nm. Reduced hemoglobin absorbs light at 660 nm. It is possible to place sensors that will determine the concentration of oxygenated and reduced, and thus total hemoglobin in the blood.(8,9) The pulsatile arterial signals are filtered from non-pulsatile components (such as bone) to calculate the concentrations of the different spectrophotometric hemoglobins, and thus the device can estimate the total concentration of all hemoglobin species combined.

There are two main types of device. One uses spectrophotometry and the other uses photoplethysmography. Both use light absorption, but different sensors and algorithms. They have a bias (difference from the autoanalyzer, the gold standard) of about 0.1 to 1.1 g/dl. Spectrophotometry devices are slightly more accurate, but the photoplethysmography devices are cheaper, smaller and lighter.(10–14) While noninvasive testing does not replace laboratory blood tests, it is highly valued for its ability to show real-time trends, helping clinicians catch sudden drops in Hb faster than intermittent blood draws. Accuracy may decrease during states of low perfusion, or with persons who have very dark skin tones.(15–18)

The Vivaray Hb Pro device is a photoplethysmography instrument. It is a quantitative, non-invasive device for screening and on-the-spot classification of hemoglobin levels among children (greater than 8 years) and adults in outpatient clinics, primary health setups, healthcare camps and blood transfusion centers. It is not intended for use at home.

The result from Vivaray Hb pro is both quantitative and qualitative. The quantitative values for hemoglobin are shown in grams per deciliter. The qualitative results are displayed as normal, mild anemia, moderate anemia, and severe anemia (Table 1).

**Table 1.** Definitions of mild, moderate and severe anemia.

| Severity of anemia | Hemoglobin level (g/dl) |  |  |
| --- | --- | --- | --- |
| | Female | Male | Child ($\geq 8$ years) |
| Severe | < 8 | < 8 | < 8 |
| Moderate | 8–10.9 | 8–10.9 | 8–10.9 |
| Mild | 11–11.9 | 11 – 12.9 | 11–11.9 |
| Normal | $\geq 12$ | $\geq 13$ | $\geq 12$ |

## Objectives

The purpose of this study is to validate the clinical performance of Vivaray Hb pro and use the device generated hemoglobin value for validation by comparing it with the hemoglobin value generated from the gold standard (calibrated blood cell counter). Specifically, we intended to measure the agreement between values obtained by using this noninvasive device with venous samples measured on a calibrated blood cell counter.

## Methods

Subjects 8 years and older were prospectively recruited from the outpatient departments of the City X-ray and Scan Clinic, Dr Dipti’s Multispeciality Clinics, and Panchsheel Hospital, New Delhi, and from Citizen Hospital, Bangalore. The convenience-based recruitment started on 13-Jun-2025_and concluded on 13-Aug-2025. All the participants provided written informed consent prior to participation in the study. The informed consent process was conducted by the principal investigator, and signed consent forms were obtained before performing any study-specific activities. A copy of the signed informed consent form was provided to each participant and the original was retained in the respective participant file. For participants who was minor, written informed consent was obtained from their parents before any study-related procedures were conducted. Assent was also obtained from participants who were minor. Copies of both signed consents were provided to the participant and parent and the originals were retained in the respective participant file.

The Vivaray Hb pro device works on the principle of ‘opto-electromechanical spectrometry’. The device utilizes specific wavelengths of light to detect photoplethysmograms (PPGs) on a human finger without pricking the skin. All centers had the calibrated cell counters working on photometry (transmittance) principles. The device is designed to fit fingers 8 to 23 mm in thickness. The inclusion criteria were finger size 8-23 mm, age above 8 years, and hemoglobin between 4 and 18 g/dl in the sample obtained by venipuncture. A single reading was obtained per participant during the study. Figure 1 shows the flow of participants.

**Figure 1.**
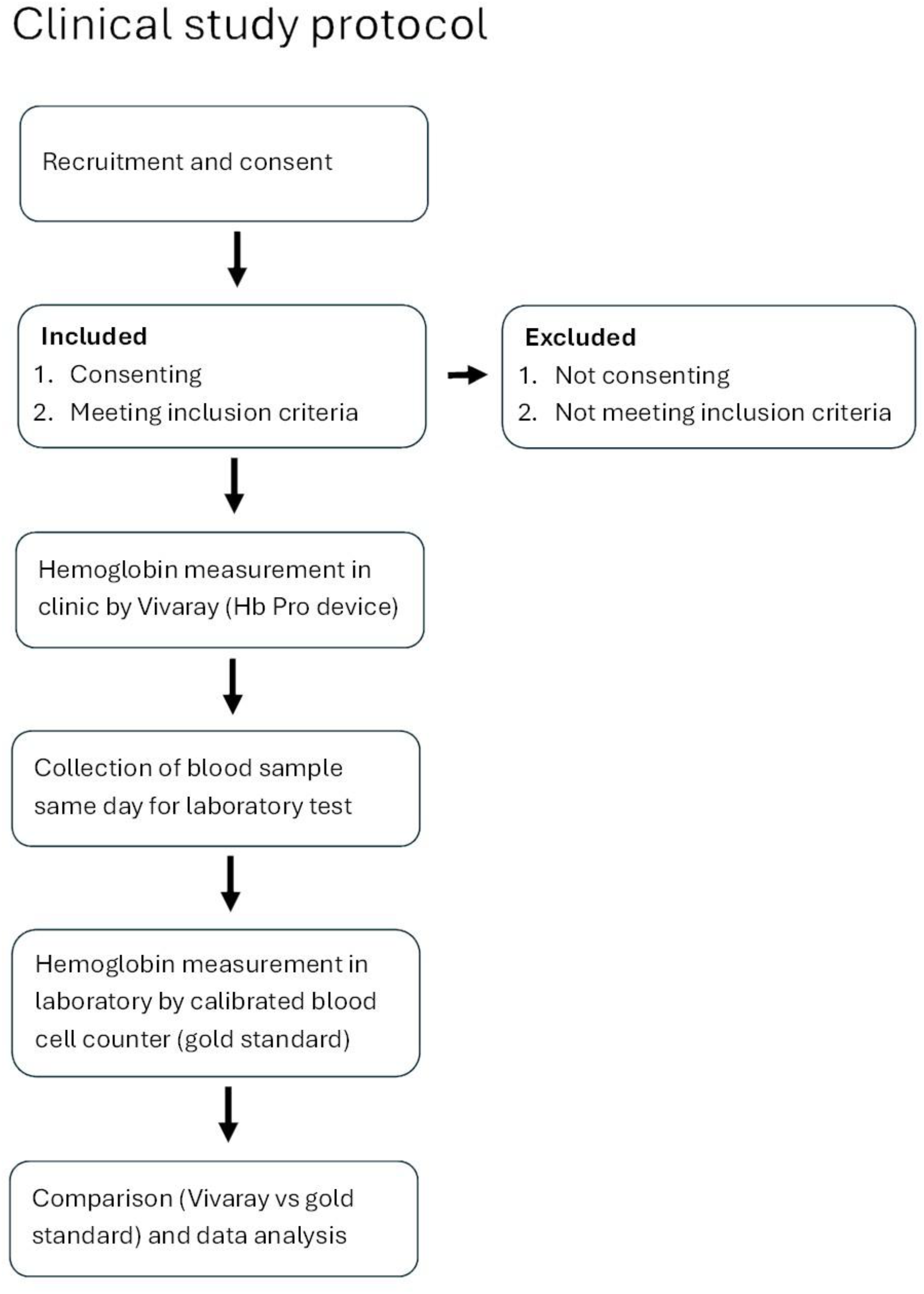
Flow chart showing subject movement and sample collection.

We excluded patients with dermatological diseases of the fingers, as well as those whose fingers were stained with nail polish or henna.

The index finger was preferred for noninvasive measurement. If the index finger was not suitable, another finger was used. Participants were instructed to place the index finger inside the finger slot, ensuring that the fingertip touched the stopper. The scan button was pressed for two seconds to begin the scanning process, which took 30 to 40 seconds for for the measurement to be complete. At the same time blood was drawn and sent to the pathology laboratory for estimation of hemoglobin levels on a calibrated cell counter.

### Statistics

The sample size was calculated for a Bland-Altman analysis. We used Medcalc software to estimate sample size with the following presumptions based on our pilot tests: alpha = 0.05, beta = 0.2, expected mean of differences −0.25, expected standard deviation of differences 1.42, and maximum allowed difference between methods = 3.23. This provided a sample size of 763 subjects. Once the data was available, the reference (Coulter) was compared with the test machine (Vivaray). Agreement was estimated by the Bland-Altman method.(19,20) Calculations were carried out using R. Subjects with missing values were excluded.

### Ethics approval

The conduct of this study adhered to the rigorous ethical and regulatory standards. Ethics approvals were obtained from the following Institutional Ethics Committees: Good Society of Ethical Research (Citi X-Ray and Scan Clinic, 29 June 2024 and Dr. Dipti’s Multispeciality Clinics, 29 June 2024), Ethics Committee Panchsheel Hospital (Panchsheel Hospital, 23 July 2024), and Citizen Hospital Institutional Ethics Committee (Citizen Hospital, 29 June 2024). The study was registered with Clinical Trials Registry, India (CTRI) having the registration number: CTRI/2024/07/071199 dated 24-Jul-2024

### Reporting

This report follows the STARD guidelines (Standards for the Reporting of Diagnostic Accuracy studies).(21)

## Results

The study commenced with the recruitment of the first participant 13-Jun-2025. The last participant was measured on 13-Aug-2025. In all, 765 participants were recruited. The noninvasive hemoglobin values and the laboratory values were recorded in all cases except two, leaving 763 participants for complete analysis. The mean age was 38 years (range 17-82 years, and 81% of participants were female (Table 2)

**Table 2.** Demographic details of the participants.

| Parameter | (N=763) |
| --- | --- |
| Age (Years) | 37.99 (s.d. 13.15) |
| Gender | Female 620 (81.3%),<br>Male 143 (18.7%) |
| Body Mass Index (BMI) (kg/m<sup>2</sup>) | 25.64 ± 4.24 |

### Agreement

Agreement was measured by a Bland-Altman analysis. The overall mean difference (test minus Coulter) was −0.40 g/dl. The mean absolute error was 1.53 g/dl.

The mean difference varied at different hemoglobin levels, overestimating the true hemoglobin at low levels and underestimating it at high levels. Figure 2 plots the true hemoglobin levels against the difference for individual readings.

**Figure 2.**
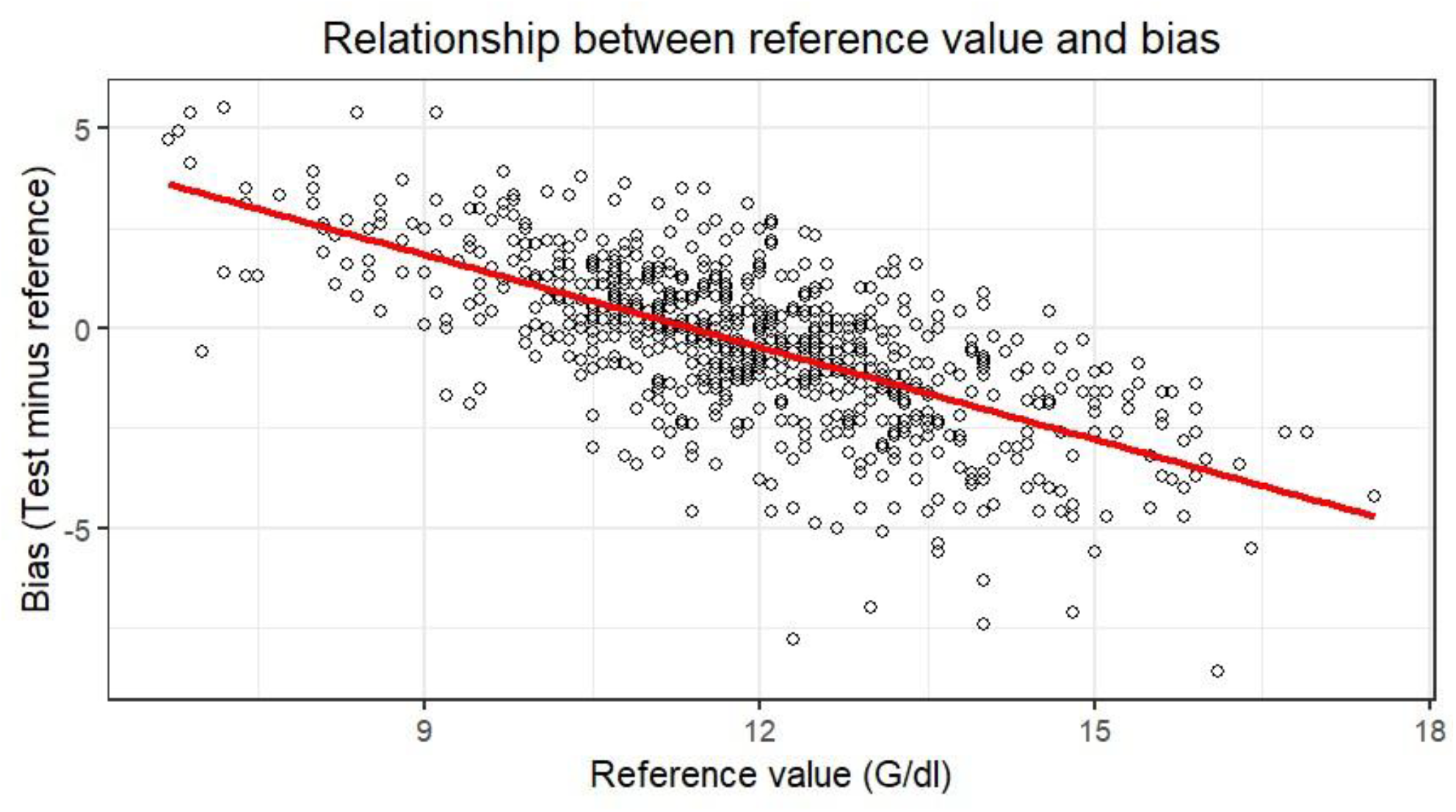
Bias at different hemoglobin levels.

Since the mean difference varied at different hemoglobin levels, we carried out a regression-based Bland-Altman analysis (Figure 3) The intercept (β_0_) was 1.68, and the regression slope (β_1_) was −0.178 (Fig 2). The regression-based Bland–Altman analysis showed proportional bias. Its slope (β_1_) was −0.178, meaning that for every for every 1 g/dL increase in the mean hemoglobin, the difference (reference minus test) changed by −0.178 g/dL. The clinically relevant bias varies with the measurement level according to the equation “bias(x) = b0 + b1x”. The test overestimated hemoglobin concentrations at low hemoglobin levels, and underestimated the concentrations at high levels.

**Figure 3.**
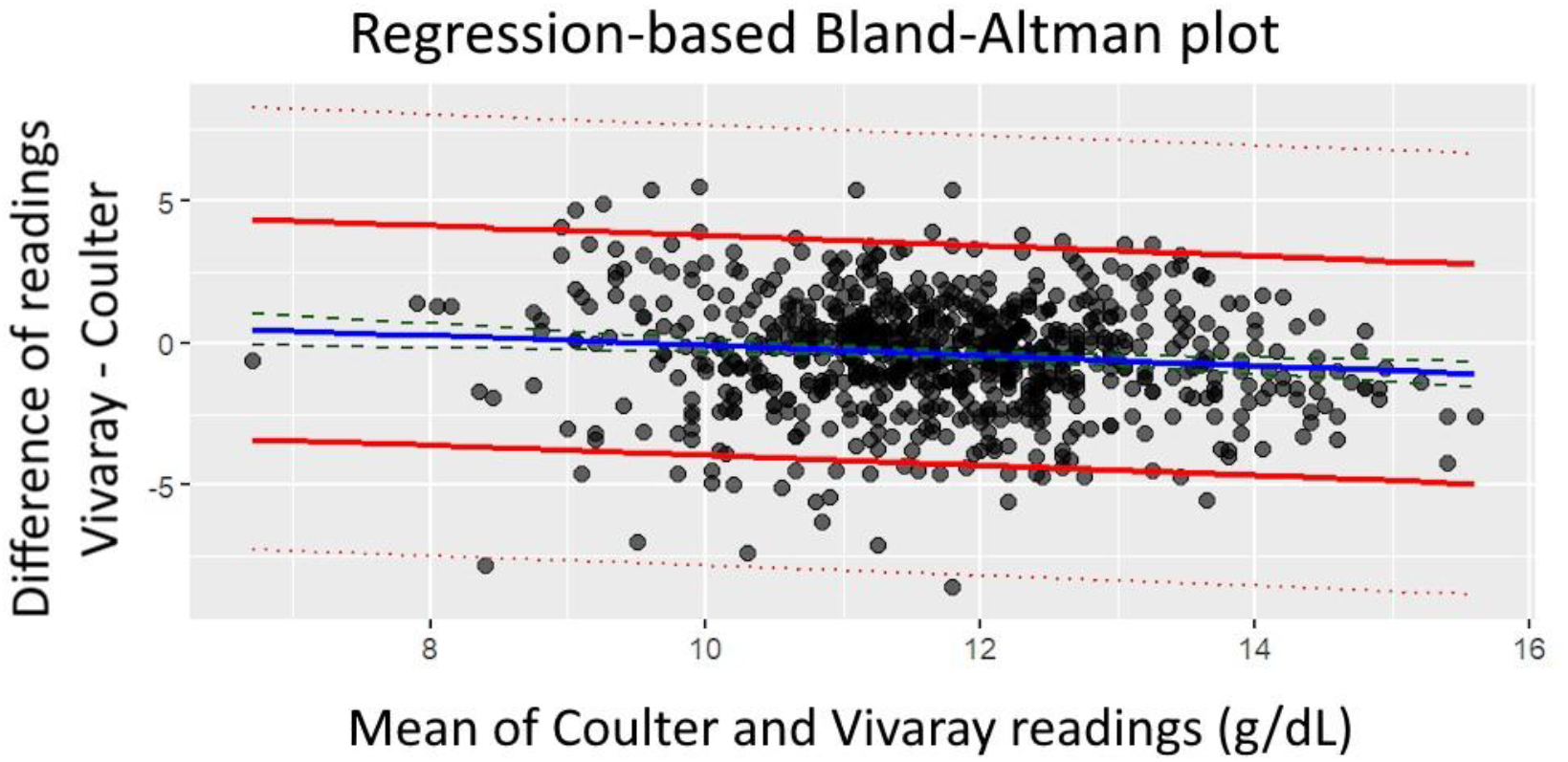
Regression-based Bland–Altman Plot: Bosch Device vs Coulter Reference. In this figure, the solid blue line represents the fitted regression line, and the adjacent dashed lines represent the 95% confidence limits around that regression line (the uncertainty around the regression-estimated bias). The solid red lines represent the regression-based limits of agreement, and the dotted red lines represent the 95% confidence limits for the limits of agreement. The regression intercept is 1.68 (95% confidence limits 2.96, 0.40). The intercept is numerically equal to the predicted difference at mean = 0 but can also be interpreted as the adjusted mean difference after accounting for proportional bias.

The predicted bias at hemoglobin 0, 9 and 15 is 1.68, 0.079 and −0.987 respectively (Table 3).

**Table 3.**
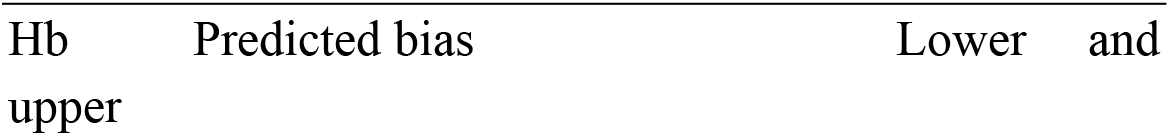
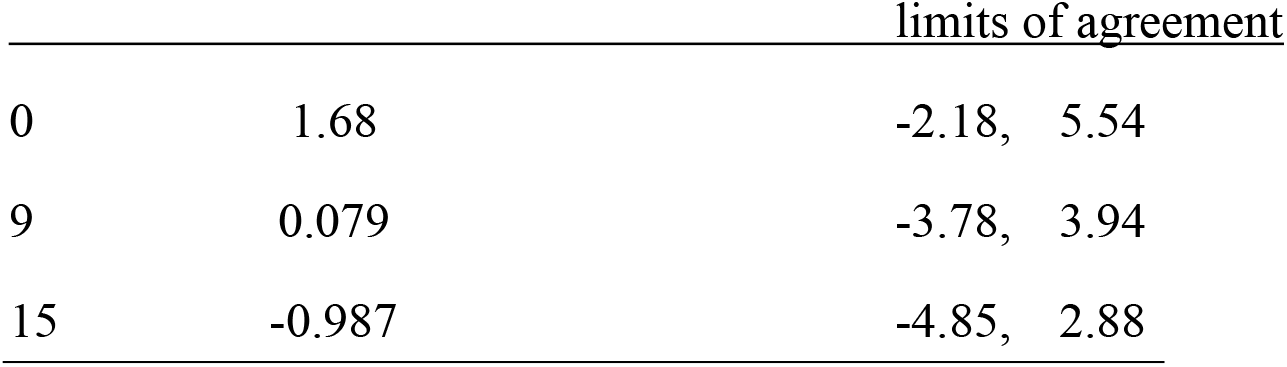
Predicted bias at different hemoglobin values.

## Discussion

It has been recognized since the early 1970s that oxygenated and deoxygenated hemoglobin absorb light differently at specific wavelengths.(22) This principle underlies modern pulse oximetry, in which a photodetector measures light at two wavelengths (660 nm and 940 nm). By separating the pulsatile arterial component from the non-pulsatile tissue signal using Aoyagi’s ratio-of-ratios method, the device estimates arterial oxygen saturation.

Non-invasive hemoglobin measurement extends this concept by using multiple wavelengths to spectrally resolve different hemoglobin species, producing a continuous estimate of total hemoglobin through pulse co-oximetry. The first clinically useful devices employing this technology were introduced by Masimo in 2008–2009.(23) Hemoglobin concentrations determined by this spectrophotometric method are usually termed SpHb to distinguish them from the total hemoglobin concentration obtained in a laboratory from venipuncture samples.

Non-invasive hemoglobin monitoring using pulse co-oximetry has demonstrated clinical utility across a broad range of care settings. It has been tested in an outpatient setting,(17,24) emergency department, (25) in neonates(26) and in intensive care units.(4,15) Noninvasive measurement significantly reduces the anxiety associated with venipuncture,(25) and seems likely to provide the benefit of continuous measurement.(4) The devices have been used often in operation theaters,(27– 32) where there seems to be a trend towards improved decision-making regarding blood transfusions.(15,27,28,31)

Nevertheless, several reports express doubts about the reliability of non-invasive devices, given the extent of the bias. (25,29,30) The concerns are that the wide limits of agreement cast doubt on the clinical usefulness of such devices. If transfusion is being considered, other testing methods may be required.

Moreira et al(33) have recently conducted a systematic review to assess the diagnostic accuracy of co-oximetry compared versus reference values in perioperative management. In 39 studies published between 2011 and 2024, they calculated the 95% limits of agreement between test and reference values. The width between the upper and lower limits represents the range within which 95% of all of the individual paired differences are likely to lie and is a measure of the precision of the test instrument. An analysis of their tables showed that the minimum width was 1.55, and the maximum width was 8.0. Moreira and coworkers interpreted their data as being supportive of the reliability of pulsed co-oximetry. To see if the precision of the machine had improved over the years we looked at the correlation between year of publication and width of the 95% confidence limits of the Bland-Altman limits of agreement in the studies (Fig 4).

**Figure 4.**
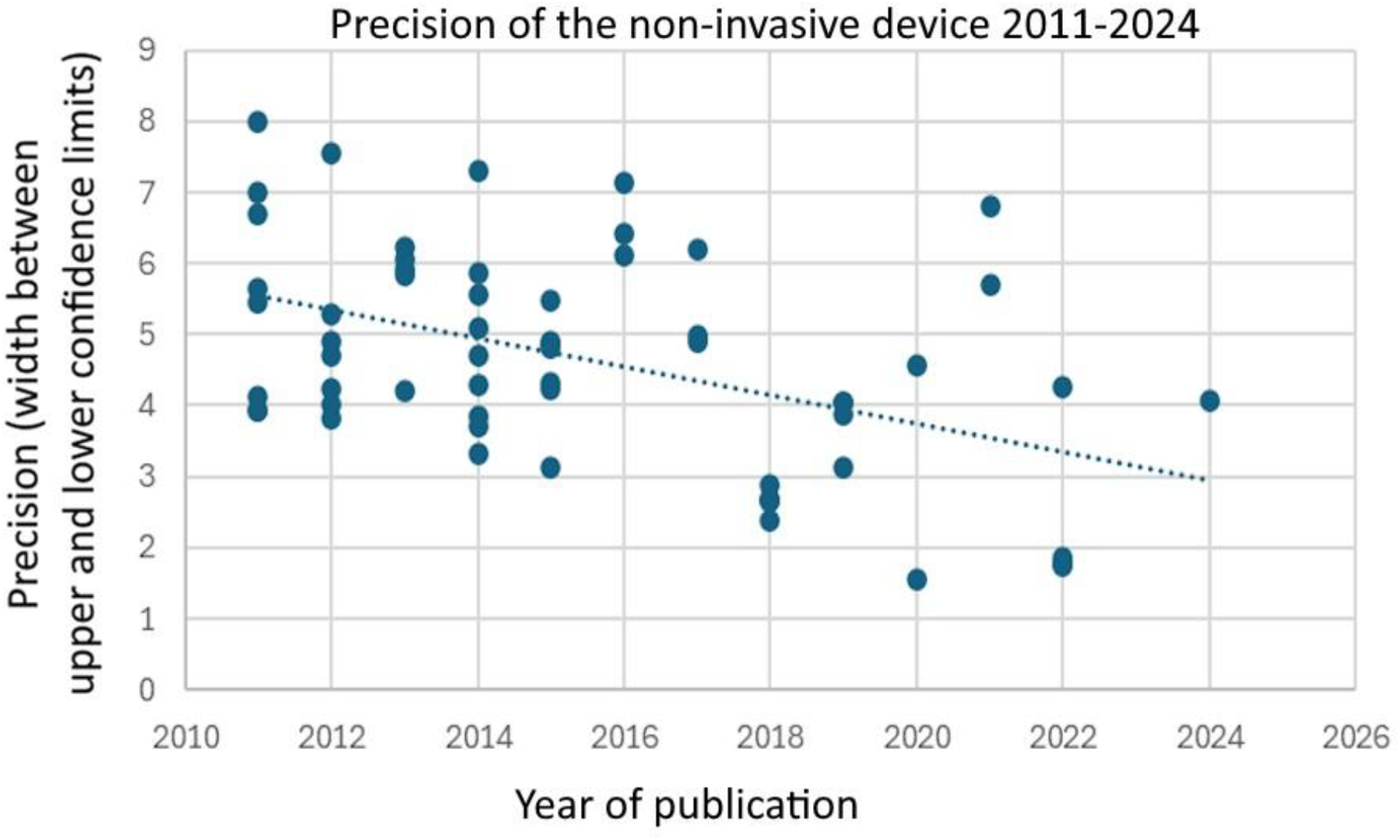
Precision of non-invasive measurement over the years. This figure is an analysis of the tables in a recent systematic review. (33)

Pearson’s coefficient of correlation was −0.46 (95% confidence limits −0.64, −0.24), indicating that since 2011 there has been a small but statistically significant improvement in the precision of the devices.

Our study shows a performance similar to that seen by other workers.(33) The bias varies at different hemoglobin levels, as seen in Fig 2 and Table 3 above, showing a phenomenon commonly seen with non-invasive devices, that of a “regression to the mean” bias or proportional bias. In the clinically predominant range (8.1-13 g/dl) the absolute mean bias was 1.25 g/dl. This, and other devices typically overestimate low hemoglobin values and underestimate high values.(15,18,34,35)

## Conclusions

This large, prospective, cross-sectional clinical performance study demonstrates that the Vivaray Hb pro is capable of providing noninvasive hemoglobin estimates with good agreement when compared with a calibrated laboratory blood cell counter. The device showed an overall mean absolute error of 1.53 g/dL, but a mean absolute error of only 1.25 g/dl in the clinically predominant range of 8.1 to 13 g/dl. Most measurements fall within clinically relevant tolerance limits. The observed pattern of hemoglobin dependent bias-characterized by overestimation at lower hemoglobin levels and underestimation at higher levels-is consistent with the behavior reported for other noninvasive hemoglobin technologies. Importantly, these findings confirm that the performance of the Vivaray Hb pro is comparable to that of existing photoplethysmography based devices used in clinical practice.

Taken together, the results support the use of the Vivaray Hb pro as a noninvasive hemoglobin screening and point-of-care assessment tool, particularly in settings where rapid, painless, and repeatable measurements are desirable. While laboratory hemoglobin estimation remains essential for definitive diagnosis and clinical decision-making, the Vivaray Hb pro offers a practical solution for initial screening, on the spot classification, and in outpatient clinics, primary healthcare facilities, and community screening programs. The device therefore represents a valuable adjunct to conventional testing methods in efforts to improve anemia screening and early identification in resource constrained and high-throughput environments.

## Strengths and weaknesses

A key strength of this investigation is the large sample size of 763 participants, making it one of the larger real-world evaluations of a noninvasive hemoglobin device conducted in outpatient and community healthcare settings. The use of Bland–Altman analysis, complemented by a regression-based approach to account for proportional bias, provides a robust and transparent assessment of agreement across the hemoglobin range. The prospective data collection, simultaneous noninvasive and venous measurements, and inclusion of participants across a wide range of hemoglobin values further strengthen the reliability and generalizability of the findings.

While this study is one of its kind to have a greater sample size compared to the studies of other non-invasive hemoglobin devices published studies, the authors would like to highlight the areas of improvement for future studies. Firstly, although individuals aged eight years and older were eligible for inclusion, the study was not designed or powered to permit a dedicated subgroup analysis for pediatric populations; therefore, conclusions regarding device performance in children remain limited. Secondly, repeated measurements were not performed, precluding evaluation of within subject variability or test–retest reliability. Future studies incorporating broader demographic representation, targeted pediatric cohorts, repeated measurements, and other real-world deployment scenarios would further strengthen the evidence base and help refine the clinical role of non invasive hemoglobin screening technologies.

## Conflicts

Dr Devesh Kumar and Suneet Sood are consultants for Think-i, Unit of Tenet Health Edutech Pvt. Ltd., the Clinical Research Organization that conducted the study.

## Acknowledgments

We would like to acknowledge the efforts of the research team, nursing team and subjects who voluntarily participated in this study.

We are grateful to Mr. Kamal Shahani, Dr. Piyush Kumar Pandy, Mr. Shaitan Singh, Kr. Chinmayee Sahoo and Mr. Prince Raj for their invaluable support and contributions in the overall management and successful execution of the study.

We are grateful to Bosch Global Software Technologies Private Limited for funding the study.

## Data availability

All data files are available from the Harvard Dataverse repository database at the following url:: https://doi.org/10.7910/DVN/JCAIFK.(36)

## Author contributions

Devesh Kumar: Conceptualization, formal analysis, funding acquisition, methodology, project administration, resources, software, supervision, validation, visualization, writing – review and editing

Sunita Kapoor: Data curation, writing – review and editing

Ambanna Gowda: Data curation, Writing – review and editing

Dipti Gupta: Data curation, writing – review and editing

Harsh Mittal: Data curation, writing – review and editing

Suneet Sood: Concept, formal analysis, writing – original draft, review and editing

## Notes

### Competing Interest Statement

I have read the journal's policy and the authors of this manuscript have the following competing interests: Devesh Kumar and Suneet Sood are consultants for Think-i, Unit of Tenet Health Edutech Pvt. Ltd., the Clinical Research Organization that conducted the study.

### Clinical Trial

The study was registered with Clinical Trials Registry, India (CTRI) having the registration number: CTRI/2024/07/071199 dated 24-Jul-2024 Powered

### Funding Statement

Yes

### Author Declarations

Ethics approvals were obtained from the following Institutional Ethics Committees: Good Society of Ethical Research (Citi X-Ray and Scan Clinic, 29 June 2024 and Dr. Dipti’s Multispeciality Clinics, 29 June 2024), Ethics Committee Panchsheel Hospital (Panchsheel Hospital, 23 July 2024), and Citizen Hospital Institutional Ethics Committee (Citizen Hospital, 29 June 2024). The study was registered with Clinical Trials Registry, India (CTRI) having the registration number: CTRI/2024/07/071199 dated 24-Jul-2024 Powered

